# The potential of CYP2D6 phenotyping to improve opioid dosing strategies

**DOI:** 10.1101/2021.10.07.21264057

**Authors:** Ridley Cassidy, Ewan Main

## Abstract

**Background:** Prescription opioids account for more than 40% of opioid induced mortalities. With the trend towards increased opioid prescribing set to continue rising, opioid-based adverse drug reactions, (ADRs) and their associated human and financial cost present a major global public health concern. The review examined the relationship between CYP2D6 phenotypes and opioid metabolising. The aim was to establish whether screening for CYP2D6 phenotypes would improve existing opioid dosing strategies and reduce ADRs.

**Method:** A systematic review was conducted using the online Web of Science database. Selected key words and Boolean operator combinations were used to search the relevant literature. Identified studies were screened against pre-defined inclusion/exclusion criteria. Eligible studies were subject to full review and quality assessment. A narrative analysis was performed to synthesise data from the studies included in the final review.

**Results:** The review yielded five studies that met the eligibility criteria and were subject to full review. Four of the five studies reported significant effects of CYP2D6 phenotypes on opioid metabolising or opioid based ADRs. Three studies focused exclusively on pharmacokinetics, two studies focused exclusively on ADRs, and one study considered pharmacokinetics and ADRs. All pharmacokinetic studies reported a significant association between CYP2D6 phenotypes and opioid metabolising. Only one of the studies reported a significant association between CYP2D6 phenotypes and ADRs.

**Conclusion:** The majority of evidence considered in the review supports the role of CYP2D6 in the metabolising of opioids and opioid based ADRs. Consequently, CYP2D6 screening should be considered as a potential mechanism for improving existing opioid dosing strategies and reducing ADRs. There is a need for further higher quality primary data studies focusing specifically on CYP2D6 phenotyping in the context of dosage strategies and exploring impact in longitudinal designs. Future studies should also seek to develop cost effective CYP2D6 screening methods to help support the clinical significance of CYP2D6 phenotyping.

## 1. Introduction

The number of drug overdose deaths has quadrupled since 1999 and is continuing to rise. Opioids account for approximately 70 percent of these deaths, with nearly half a million opioid related deaths in the last 20 years (Centre for Disease Control and Protection, 2021). Prescription opioids are reported to account for more than 40 percent of opioid induced mortalities, with this figure set to increase further as a result of an exponential rise in the number of opioid dependent patients (see figure 5) (Schnell, 2019).

Adverse drug reactions (ADRs) are a growing cause of mortality, affecting quality of life, increasing demand on healthcare systems and presenting a major public health concern (Khalil & Huang, 2020). ADRs account for 6.5% of total hospital admissions in the United Kingdom at a projected annual cost of £466 million (Pirmohamed et al., 2004). Pharmacokinetics relates to the absorption, distribution, metabolism and excretion of a drug in the body (Ratain & Plunkett, 2003). Patient heterogeneity in drug response can lead to disruption of the pharmacokinetics of opioids, resulting in ADRs and reduced overall therapeutic efficacy. One aspect of this heterogeneity relates to genetic interindividual variability affecting the rate at which individuals metabolise opioids. Ultrarapid metabolisers ‘convert’ opioids faster than poor metabolisers.

The significance and impact of this interindividual variability is illustrated in a statement issued by the US Food and Drug Administration (FDA):

> Respiratory depression and death have occurred in children who received codeine in the post-operative period following tonsillectomy and/or adenoidectomy and had evidence of being ultra-rapid metabolizers of codeine (i.e., multiple copies of the gene for cytochrome P450 isoenzyme 2D6 [CYP2D6] or high morphine concentrations). Deaths have also occurred in nursing infants who were exposed to high levels of morphine in breast milk because their mothers were ultra-rapid metabolizers of codeine. (FDA, 2015)

Such cases, together with cases of poor metabolisers who may experience inadequate pain relief (Dean, 2017), highlights the potentially critical role of genetic interindividual variability in the outcome of opioid treatment. The role of the cytochrome P450 (CYP450) enzymatic family, and CYP2D6 in particular, in the metabolism of prescription opioids may provide valuable insight helping to refine the therapeutic window and improve dosing strategies, reducing opioid related ADRs and associated human and financial cost.

### 1.1 Opioids and the Therapeutic Window

The parameters governing pharmaceutically relevant analgesia versus potentially fatal respiratory suppressive activity of opioids is referred to as the *therapeutic window*. Commonly defined as the concentration of drug quantities at which the drug is effective without activation of ADRs, the therapeutic window is determined using the therapeutic index (median lethal dose (LD_50_ / median effective dose ED_50_). In the case of opioid-like drug groups, where the therapeutic window is narrow, determining accurate dosage levels is critical to reducing the risk of ADRs. The intense analgesic and respiratory suppressive nature of opioids is a function of the activation of the mu opioid receptor (MOR), a G-Protein Coupled Receptor (GPCR). It is the interaction between MOR and βarrestin_2_ that is considered responsible for the activation of the opioids’ potentially fatal respiratory suppressive nature (Schmid, Kennedy & Ross, 2017).

### 1.2 Determining Opioid Dosage

The underlying principle governing opioid dosing strategies is to achieve and maintain rapid analgesia whilst limiting any risk to the patient (Davis, 2004). Current strategies, developed during clinical drug development trials and implemented in clinical practice, rely on a combination of basic quantitative demographic and physiological parameters (gender, weight, height and age) and more subjective and qualitative factors, including self-report pain scores. This approach to determining effective and safe dosage has been questioned, with evidence suggesting that patient weight, for example, is not reliably associated with analgesic response to opioids (Patanwala, Keim & Erstad, 2012) and populations used in clinical drug development trials fail to represent shifting population distributions around obesity, resulting in flawed weight-based pharmacokinetic models (Pai, 2012). An over reliance on qualitative and subjective patient data in clinical practice may also contribute to ineffective or unsafe dosage. Pain assessment is, understandably, a common feature in determining opioid dosage in clinical practice. While pain scores provide valuable insight into the patient’s perception of their pain and enable the patient’s response to analgesic therapy to be monitored over time, they remain inherently subjective in nature with considerable variation in the range of potential pain rating scales, including numerical, categorical and visual response scales (see Figure 2) (Schneider, Yale & Larson, 2003). This may lead to practitioner variability in interpretation and application in the context of prescribed opioid dosing. The issue may be further compounded by subjective questions such as ‘what is the worst it [pain] has been in the past 24 hours’, ‘what percentage of the time when you are awake have you had pain?’ and questions assessing functional improvement such as ‘lying in bed all day’ versus ‘very active and able to do anything you want’ (Schneider et al., 2003). Although this information could be valuable for monitoring pain during treatment, it may lack validity as an assessment for determining initial dosage.

**Figure 1.**
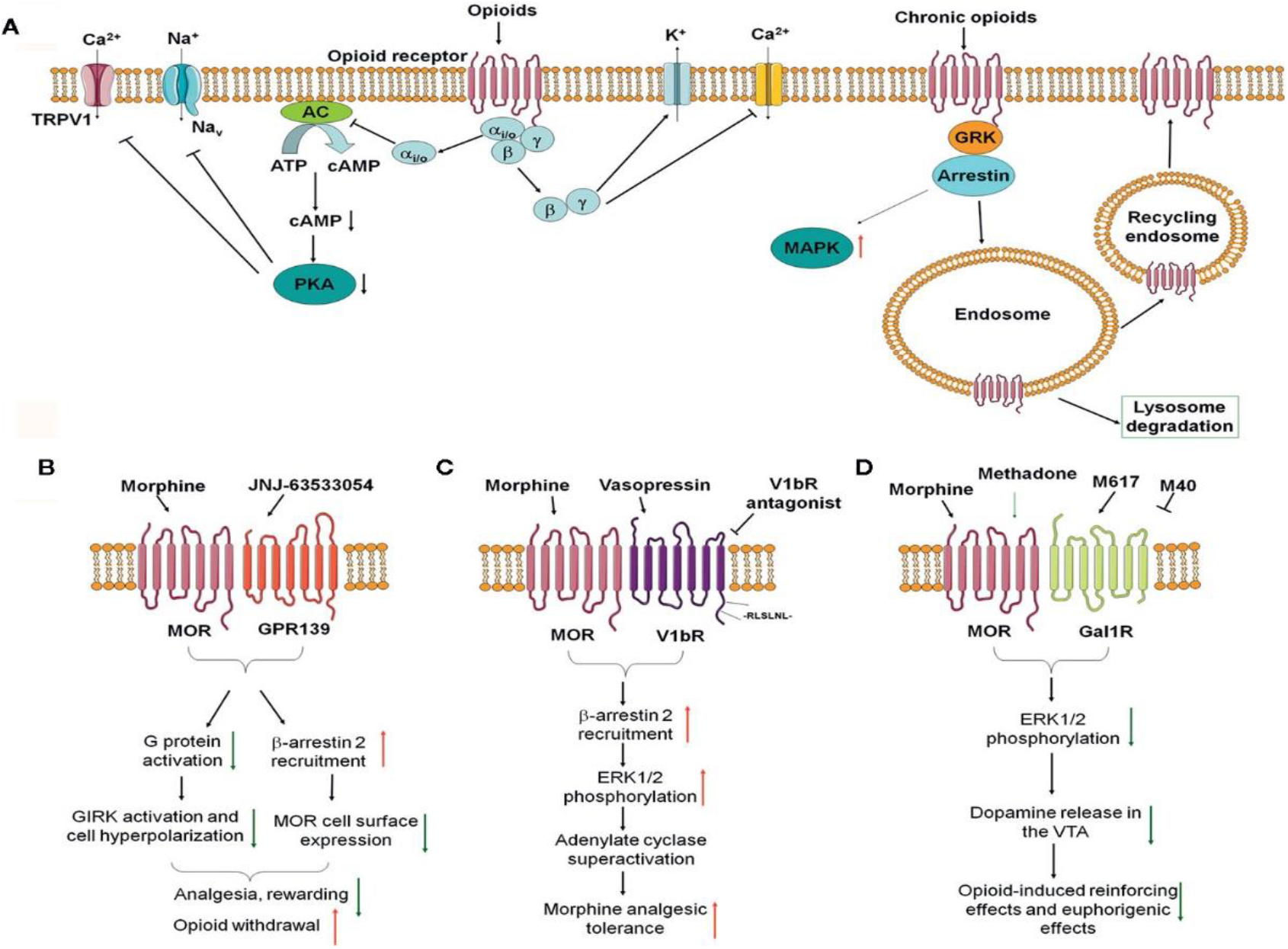
*Opioid Receptor Signaling*. (Zhang et al., 2020) *(A) Classical opioid receptor signaling pathways. (B–D) Recent identified opioid receptor heterodimers and downstream signaling in the nervous system and their effects on opioid side effects, including MOR-GPR139 (B), MOR-V1bR (C), and MOR-GalR1 (D)* (Zhang et al., 2020).

**Figure 2.**
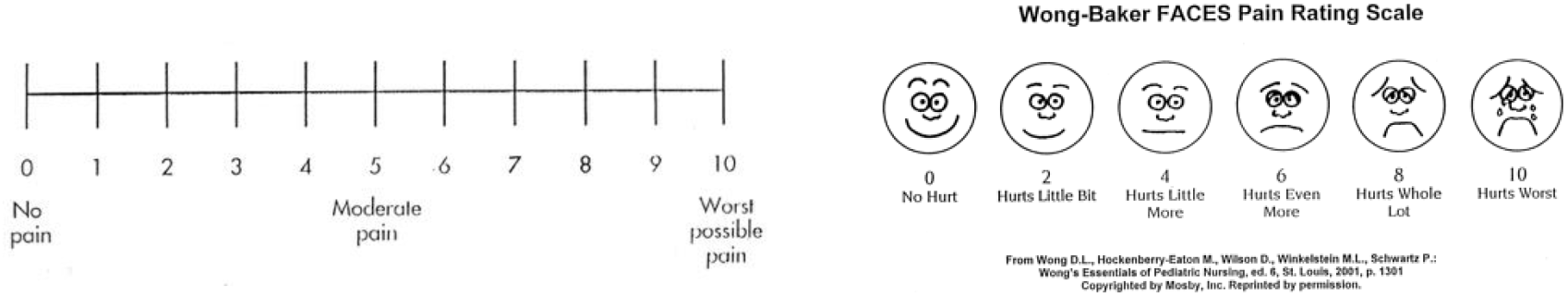
*Example Pain rating scales* (Schneider et al., 2003)

Major differences in the prescribing behaviour of doctors have also been identified, with some doctors prescribing opioids 55 times more frequently than their colleagues (Desveaux, Saragosa & Kithulegoda, 2019). Behavioural determinants, including doctors’ beliefs about capabilities, confidence, professional role, identity and emotion helped explain variations in opioid prescribing (Desveaux et al., 2019).

Taken together, these limitations raise considerable doubt regarding the efficacy of existing approaches to the development and implementation of opioid dosing strategies, highlighting potentially critical shortcomings and underlining existing calls for the reappraisal of opioid dosing paradigms (Pai, 2012). One promising yet relatively under explored avenue of investigation with the potential to further inform and refine opioid dosage strategies is genetic interindividual variability screening.

### 1.3 CYP450 Genetic Interindividual Variability in Opioid Metabolism

The central role played by cytochrome P450 (CYP450) enzymes in the metabolic pathways of opioids may offer further insight to improve opioid dosing strategies and their clinical application, improving opioid treatment outcomes. The family of CYP450 monooxygenase enzymes, found embedded in the lumen of the smooth endoplasmic reticulum, are responsible for the endogenous metabolism of around 75% of all drugs (Gregory, 2009). Although there are more than fifty known CYP450 enzymes, CYP1A2, CYP2C9, CYP2D6, CYP3A4 and CYP3A5 enzymes account for the metabolism of up to 90% of opioids and are essential in the primary metabolism of codeine, fentanyl, methadone, oxycodone, and oxymorphone (see Table 1). Moreover, most prescription opioids (codeine, hydrocodone, methadone, tramadol and oxycodone) undergo extensive dealkylation reactions (N-demethylation and O-demethylation) by CYP2D6, while a small number of highly lipophilic opioids like fentanyl (palliative care) undergo dealkylation reactions by CYP3A4 (Gregory, 2009). These type I modification reactions by CYP450 enzymes are pivotal in the metabolic activation of opioids to their highly analgesic metabolites.

**Table 1.**
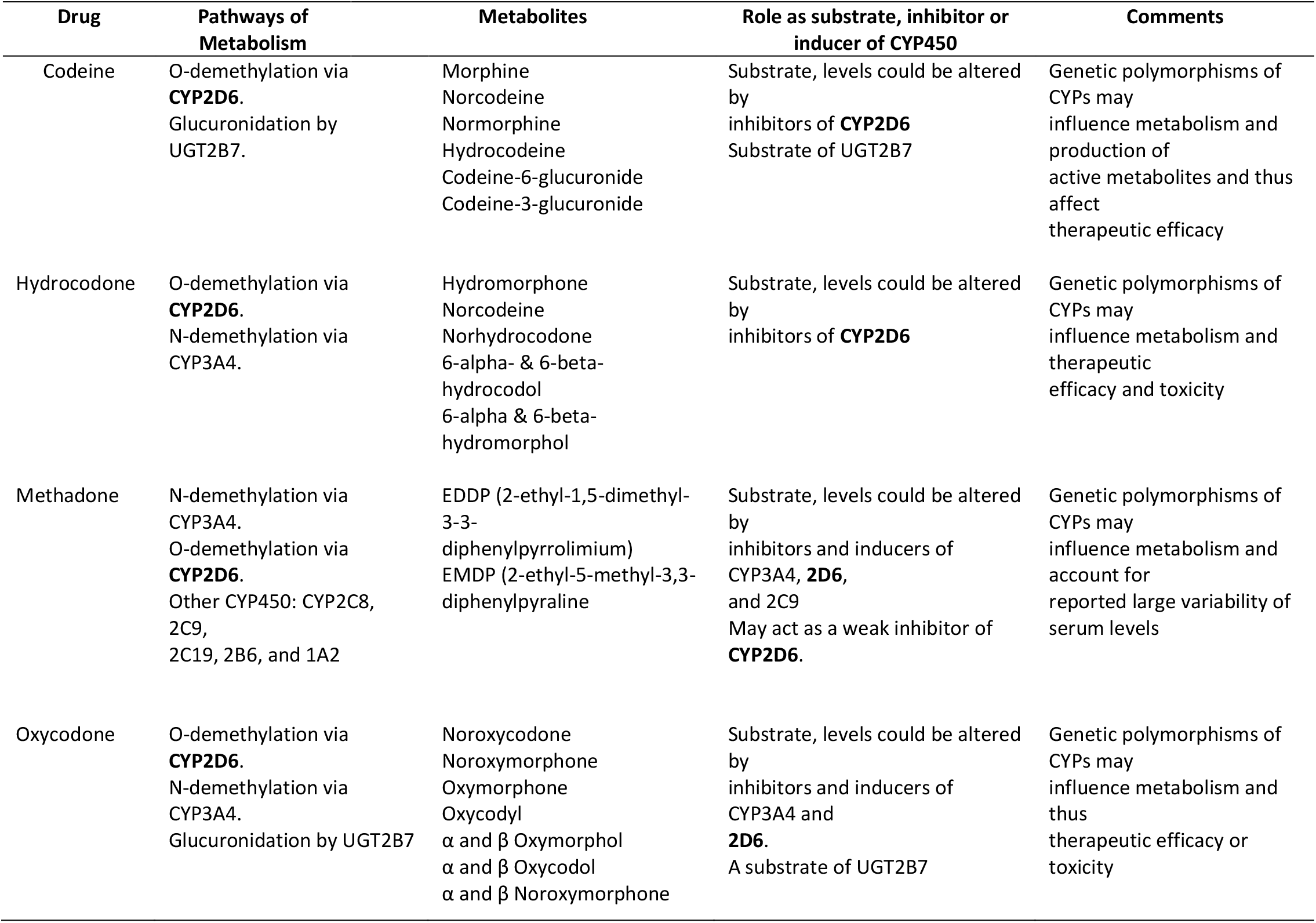
*The role of CYP450 in opioid metabolism* (Gregory, 2009)

| Drug | Pathways of Metabolism | Metabolites | Role as substrate, inhibitor or inducer of CYP450 | Comments |
| --- | --- | --- | --- | --- |
| Codeine | O-demethylation via **CYP2D6**.<br>Glucuronidation by UGT2B7. | Morphine<br>Norcodeine<br>Normorphine<br>Hydrocodeine<br>Codeine-6-glucuronide<br>Codeine-3-glucuronide | Substrate, levels could be altered by<br>inhibitors of **CYP2D6**<br>Substrate of UGT2B7 | Genetic polymorphisms of CYPs may influence metabolism and production of active metabolites and thus affect therapeutic efficacy |
| Hydrocodone | O-demethylation via **CYP2D6**.<br>N-demethylation via CYP3A4. | Hydromorphone<br>Norcodeine<br>Norhydrocodone<br>6-α- & 6-β-hydrocodol<br>6-α & 6-β-hydromorphol | Substrate, levels could be altered by<br>inhibitors of **CYP2D6** | Genetic polymorphisms of CYPs may influence metabolism and therapeutic efficacy and toxicity |
| Methadone | N-demethylation via CYP3A4.<br>O-demethylation via **CYP2D6**.<br>Other CYP450: CYP2C8, 2C9, 2C19, 2B6, and 1A2 | EDDP (2-ethyl-1,5-dimethyl-3-3-diphenylpyrrolinium)<br>EMDP (2-ethyl-5-methyl-3,3-diphenylpyraline) | Substrate, levels could be altered by<br>inhibitors and inducers of CYP3A4, **2D6**, and 2C9<br>May act as a weak inhibitor of **CYP2D6**. | Genetic polymorphisms of CYPs may influence metabolism and account for reported large variability of serum levels |
| Oxycodone | O-demethylation via **CYP2D6**.<br>N-demethylation via CYP3A4.<br>Glucuronidation by UGT2B7 | Noroxycodone<br>Noroxymorphone<br>Oxymorphone<br>Oxycodol<br>$\alpha$ and $\beta$ Oxymorphol<br>$\alpha$ and $\beta$ Oxycodol<br>$\alpha$ and $\beta$ Noroxymorphone | Substrate, levels could be altered by<br>inhibitors and inducers of CYP3A4 and **2D6**.<br>A substrate of UGT2B7 | Genetic polymorphisms of CYPs may influence metabolism and thus therapeutic efficacy or toxicity |

### 1.4 Pharmacokinetics and Mechanism of Action of CYP2D6

An understanding of the metabolic action of CYP2D6 in relation to opioids provides the basis to illustrate its potential in determining opioid dosage and guiding clinical practice in opioid treatment. The intermediate products produced as a result of metabolic action of CYP2D6 provide the highly valued analgesic metabolites of opioids (see Figure 3). The highly lipophilic nature of opioids allows for fast absorption across cell membranes. The metabolism of opioids is considered in two phases occurring mainly in the liver where they undergo first pass effect. The first phase of metabolism is defined as the modification reactions by CYP2D6 (see figure 3).

**Figure 3.**
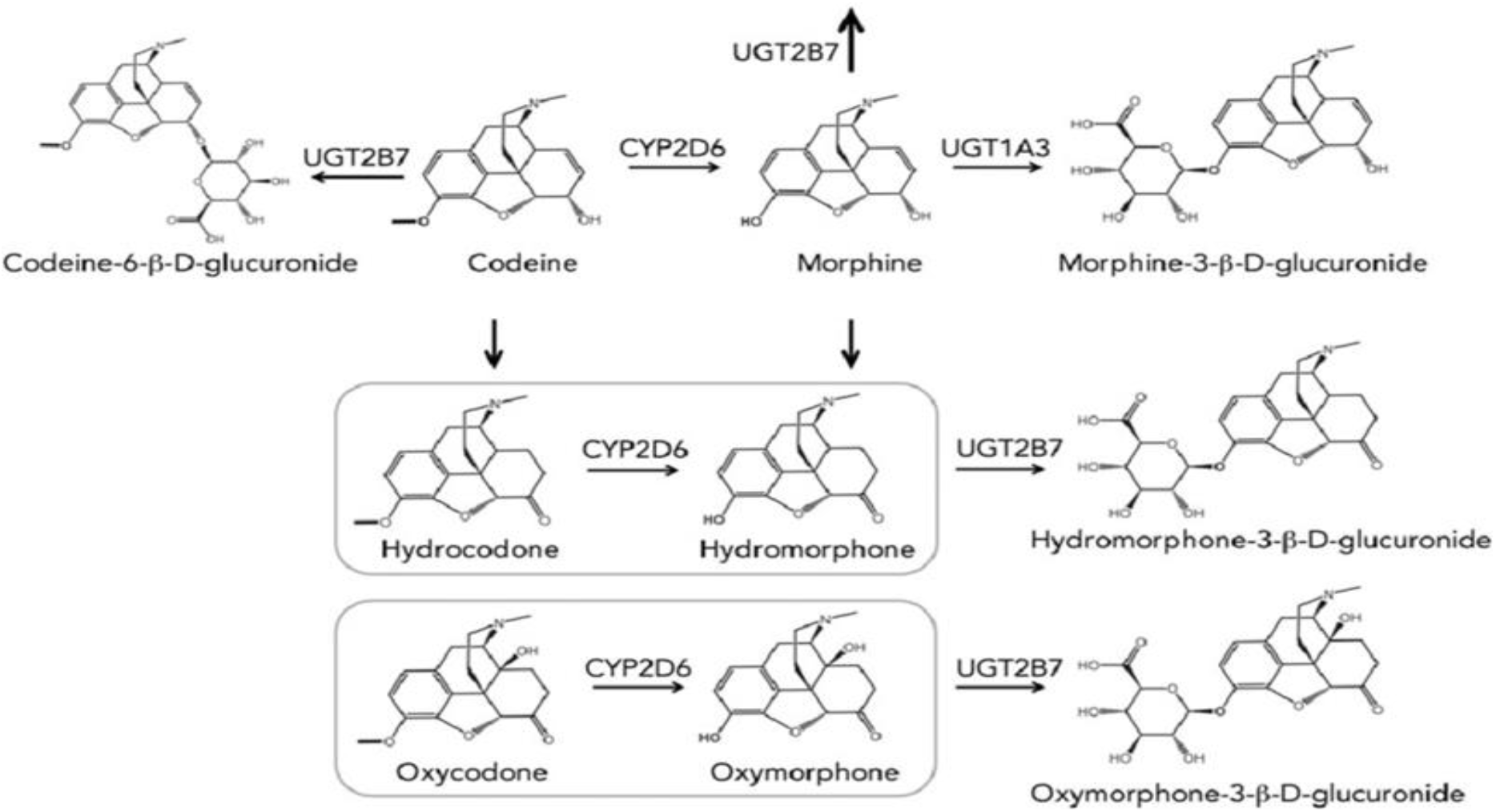
*Opioid Degradation by CYP2D6* (Marinova et al., 2016)

Perhaps the most important opioid-based metabolism is the biotransformation of codeine to its highly active analgesic metabolite, morphine by CYP2D6 (Thorn, Klein & Altman, 2009). The importance of the pharmacokinetic metabolism of codeine to morphine by variation in the CYP2D6 gene is based on morphine’s greatly increased analgesic nature, where it has an increased binding affinity of ∼200 fold for MOR and 50-times higher intrinsic activity at μ-opioid receptors, resulting from the O-demethylation of codeine, catalysed by CYP2D6 (see Figure 1) (Dean, 2017). Similar pharmacokinetic observations are made for the O-demethylation of tramadol to its active metabolite O-desmethyltramadol (ODT, M1), which expresses a ∼300-400 fold higher affinity for MOR and activity along the μ opioid receptors (Leppert, 2011). Phase I dealkylation reactions by CYP2D6 are vital in the activation of opioid metabolites.

Although all CYP450 enzymes express genetic polymorphism, CYP2D6 expresses the highest levels of interindividual variability, with more than 100 known allelic variants (Gregory, 2009). It is this degree of polymorphism that helps justify a focus on CYP2D6 over other CYP450 enzymes as a possible mechanism for improving opioid dosing strategies and reducing ADRs. The interindividual variation of CYP2D6 is considered in four phenotypic families, poor metabolisers (PM), intermediate metabolisers (IM), extensive metabolisers (EM) and ultrarapid metabolisers (UM) (Gregory, 2009). That between 77% and 92% of individuals express at least one functional metabolically active allele of CYP2D6, demonstrates high levels of CYP2D6-dependent genome interindividual variability. This is further demonstrated by inter-ethic variability data showing only 50% of CYP2D6 alleles in individuals of Asian ethnicity are metabolically active (see figure 4). These polymorphs are explained primarily in terms of single nucleotide polymorphisms (SNP). SNPs can be expressed as nonfunctional alleles, the most common of which are SNPs CYP2D6^32^ and CYP2D6^10^ (Dean, 2017). Perhaps the most critical genetic variation in the context of pharmacokinetics and opioid dosage are the 1-2% of patients who express CYP2D6 variants that result in ultrarapid metabolism (SNPs CYP2D6^1^, CYP2D6^2^, CYP2D6^17^ and CYP2D6^35^). These individuals carry at least three copies of the CYP2D6 gene and express a high level of CYP2D6 transcription (Dean, 2017) (see Table 2).

**Table 2.** *CYP2D6 alleles and their associated activity* (Fleeman et al., 2011).

| CYP2D6 Variant | Associated Enzymatic Function and CY2D6 Activity |
| --- | --- |
| *wt alleles* |  |
| *1, *2, *35 | Normal CYP2D6 Activity (*Associated with **Poor/Intermediate Metabolisers***) |
| *vt alleles* |  |
| *3, *4, *5, *6, *9 | Loss of CYP2D6 Activity (*Associated with **Poor Metabolisers***) |
| *10, *17, *41 | Decreased CYP2D6 Activity (*Associated with **Intermediate Metabolisers***) |
| *multiple alleles* |  |
| *1 x N, *2 x N | Increased CYP2D6 Activity (*Associated with **Ultra Rapid Metabolisers***) |

**Figure 4.**
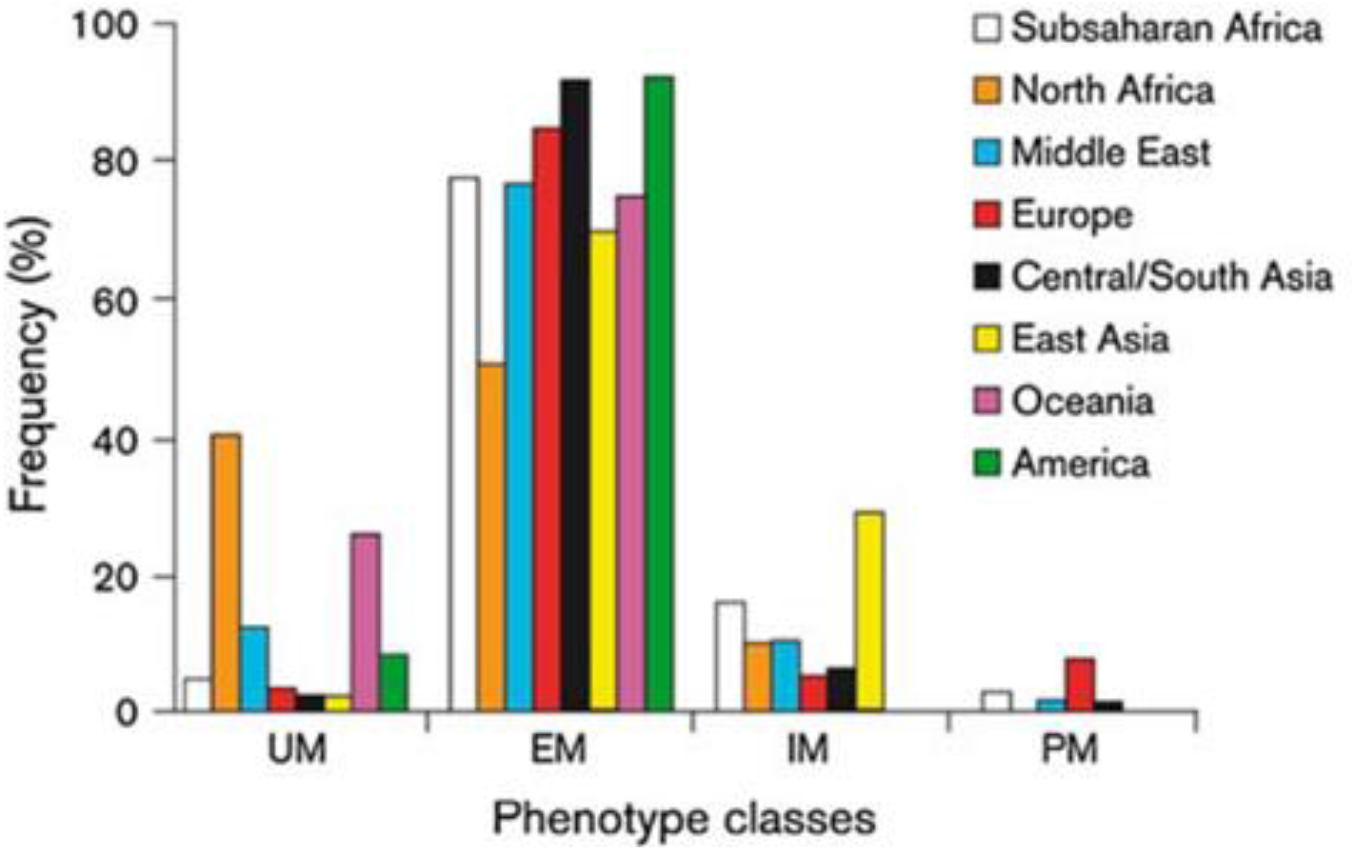
*Geographical distribution of CYP2D6 phenotype* (Karin et al., 2013).

**Figure 5.**
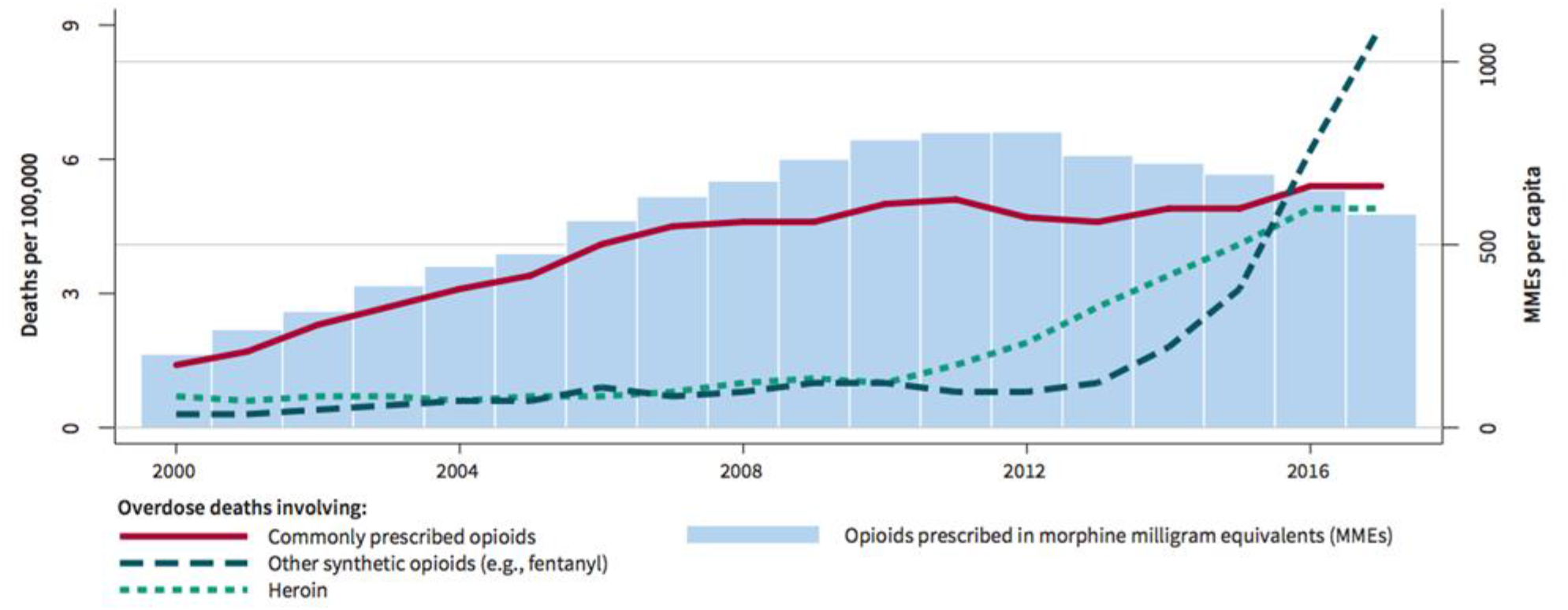
*Opioid-induced deaths between 2000 and 2017* (Schnell, 2019)

SNPs >100kb of the CYP2D6 gene are linked with significant alterations in CYP2D6 mRNA expression (Wang et al., 2014). SNPs identified in the enhancer region >100 kb downstream of CYP2D6 (rs5758550/rs133333) are found to substantially increase CYP2D6 transcription. Enhancer SNPs counteract the reduced function of SNP rs16947 in nearly half of rs16947 carrying haplotypes, while enhancer SNPs existing in the absence of rs16947 appear to convey an ultra-rapid CYP2D6 phenotype, outlining not only the significance of the specific SNPs, but also their location along the CYP2D6 gene (Wang et al., 2014).

CYP2D6 exposure to alternative splicing along two major splicing variants (SV), exon 3 (E3-SV) and exon 6 (E6-SV), causes variability in CYP2D6 gene activity. E3-SV is an in-frame deletion of exon 3, reducing the translated protein by 51 amino acids, while E6-SV encodes mRNA code lacking 142 bases of exon 6, causing an in frame shift which results in a premature termination codon 17 bases downstream (Wang et al., 2014). Alteration of exon 6 splicing by rs16947 combined with downstream SNPs in the enhancer region of the CYP2D6 has profound effect on CYP2D6 opioid metabolic activity through altered levels of transcription of CYP2D6 mRNA (Wang et al., 2014).

The prevalent role played by CYP2D6 in the metabolic pathway of many opioids is illustrated in Table 3. This, along with its highly polymorphic nature, suggests CYP2D6 as a potentially significant factor in determining safe and effective pharmaceutical interventions using opioids. Critically, exploiting CYP2D6 interindividual variability to achieve optimal individualised dosing would involve genetic screening for CYP2D6 interindividual variability.

**Table 3.** *Associated CYP2D6 substrates, inhibitors and inducers* (Mongi and Peiro, 2021)

| CYP2D6 Substrates | CYP2D6 Inhibitors | CYP2D6 Inducers |
| --- | --- | --- |
| Amitriptyline | Amiodarone | Dexamethasone |
| Amphetamine | Bupropion | Rifampicin |
| Codeine | Celecoxib | Haloperidol |
| Desipramine | Cimetidine | Glutethimide |
| Dextromethorphan | Citalopram |  |
| Donepezil | Diphenhydramine |  |
| Doxepin | Doxepin |  |
| Duloxetine | Duloxetine |  |
| Fluoxetine | Escitalopram |  |
| Haloperidol | Fluoxetine |  |
| Hydrocodone | Haloperidol |  |
| Methadone | Hydroxyzine |  |
| Metoclopramide | Methadone |  |
| Nortriptyline | Paroxetine |  |
| Oxycodone | Ritonavir |  |
| Paroxetine | Sertraline |  |
| Promethazine | Venlafaxine |  |
| Tramadol |  |  |

### 1.5 Genetic Screening for CYP2D6 Interindividual Variability

It is argued here that CYP2D6 screening to determine polymorphic variability has the potential to optimise opioid dosing, maintaining or elevating analgesia and minimising respiratory depression to reduce ADRs. Genetic testing for presence of CYP2D6 polymorphs would provide insight into the metabolic activity of the CYP2D6 in relation to phenotypic metabolisers already outlined. The genetic screening would be scored based on the alleles present, generating a quantification signifying the phenotype. This could then be factored into dosing strategies to help determine accurate opioid dosage for chronically prescribed patients, defined by the FDA as patients ‘receiving at least 60 mg of morphine daily, at least 30 mg of oral oxycodone daily, or at least 8 mg of oral hydromorphone daily or an equivalent analgesic dose of another opioid for a week or longer’ (MD Anderson Centre, 2020). Genetic screening of acute emergency medicine (ED) patients would not be practically, economically or clinically viable, hence the suggested focus on chronic prescription patients, who present the highest risk of opioid based ADRs over an extended period.

### 1.6 The Current Study

Despite the apparent potential of genetic interindividual variability to contribute to improved opioid dosing strategies, there is a paucity of available primary research specifically exploring the issue. The limited literature that is available has not been the subject of a recent review drawing together evidence in the context of pharmacokinetic and opioid dosing strategy. On that basis, the current study adopts the principles of a systematic review approach to search, screen and evaluate pertinent extant primary research to address the question of whether there is evidence to support the potential value of CYP2D6 phenotyping as a significant factor in determining opioid dosage.

## 2. Methods

No existing reviews evaluating the potential of CYP2D6 as a factor in determining accurate opioid dosing strategies were identified. The review was prepared in accordance with PRISMA (Preferred Reporting Items for Systematic Reviews and Meta-Analyses) statement which provides guidance on the minimum set of items for reporting systematic reviews (Moher, Liberati, Tetzlaff, Altmand & The PRISMA Group, 2009).

### 2.1 Eligibility Criteria

The PICOS (population, intervention, comparison, outcome and study design) framework was used to formulate the eligibility (inclusion/exclusion) criteria that determined the selection of studies to include in the review (Methley et al., 2014) (see Table 4). All population types were eligible, including studies conducted on adults and children, healthy/non-clinical and clinical samples. Studies were not excluded based on their population group due to the significance of phenotypic variability distributed among age groups and geographic locality (see Figure 4).

**Table 4.**
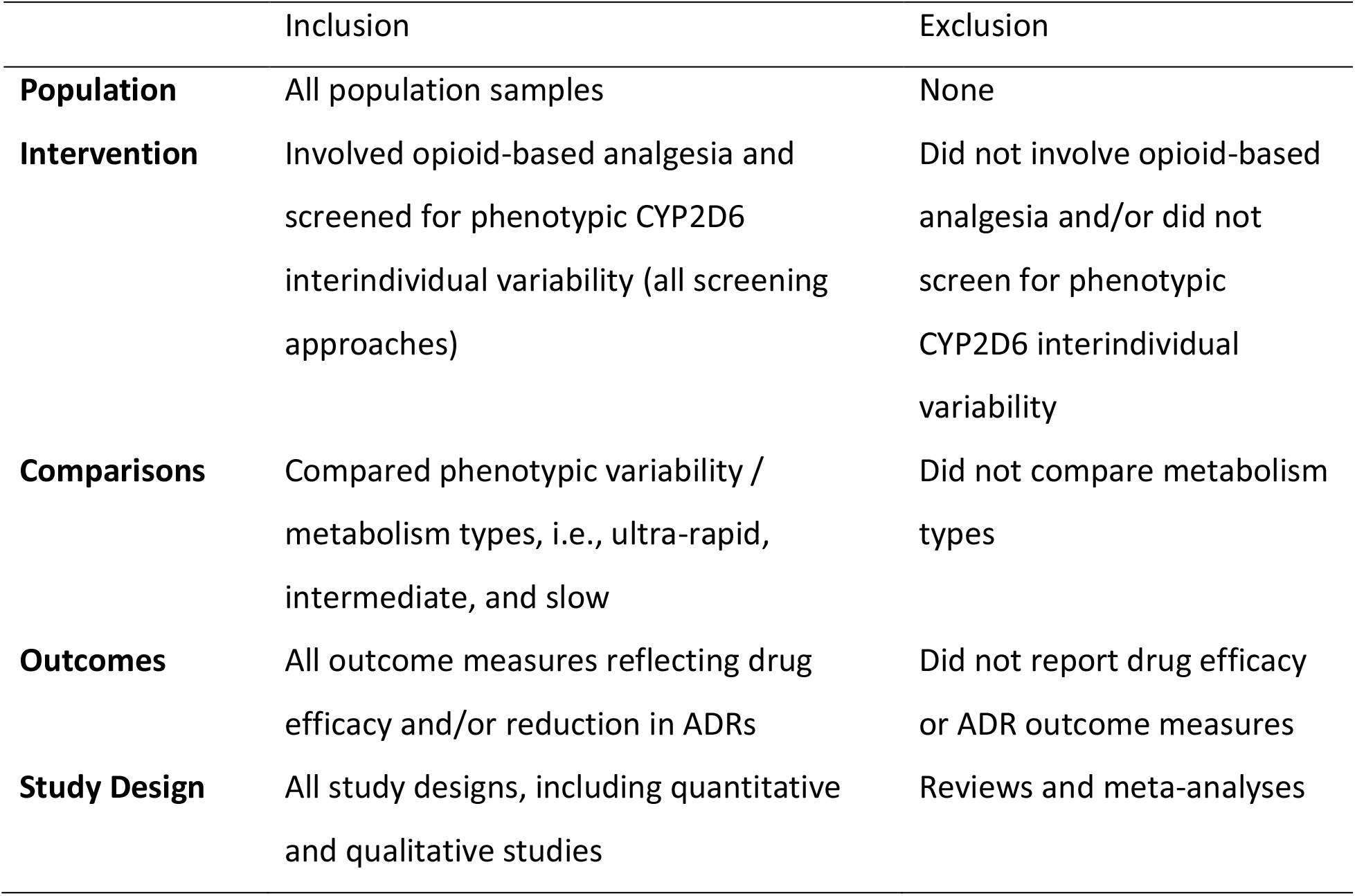
Eligibility criteria using PICOS framework

**Table 5:**
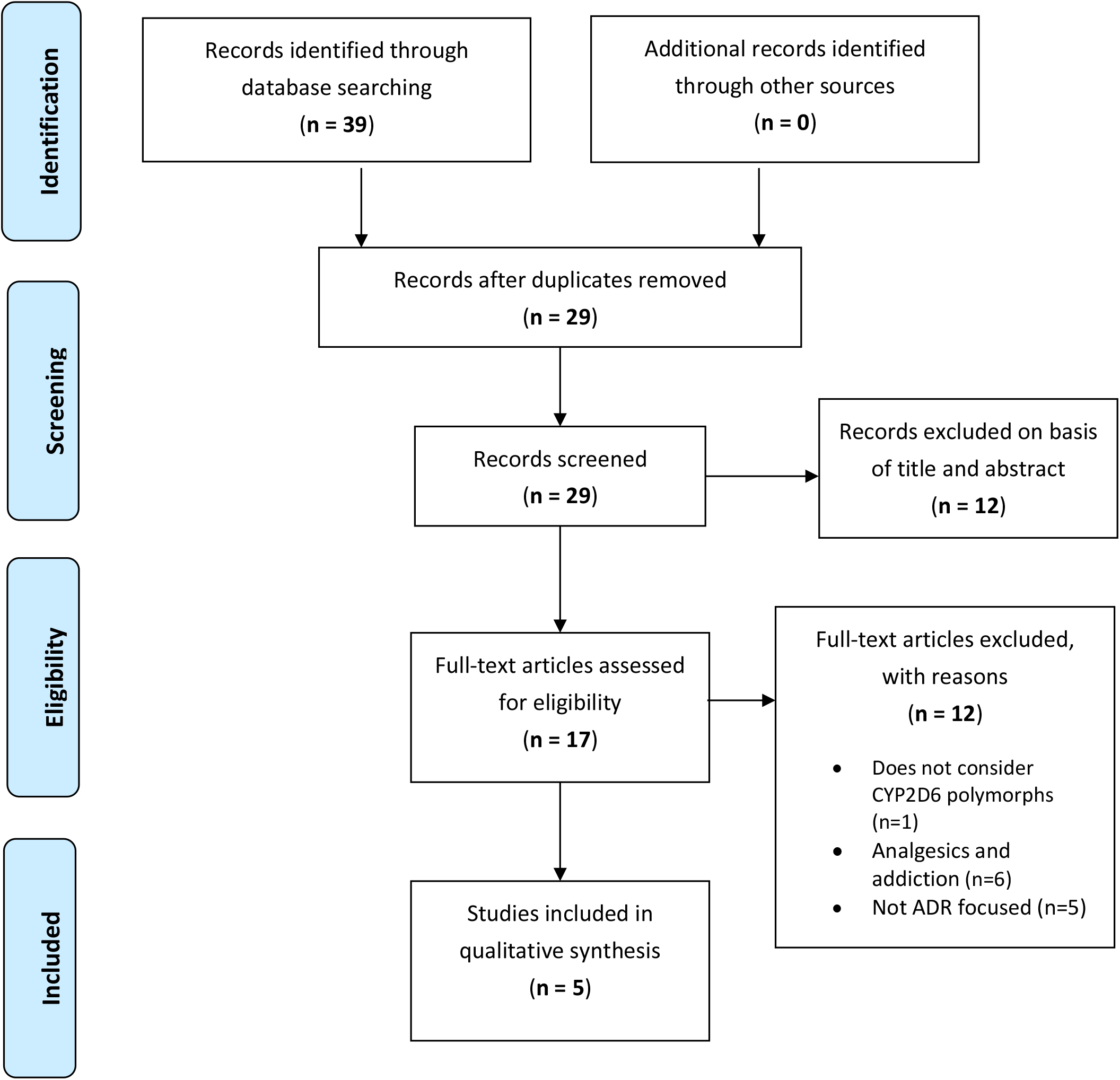
PRISMA (Preferred Reporting Items Systematic Reviews & Meta-Analyses) Flow Diagram

Interventions were restricted to those involving opioid-based analgesia and/or that screened for CYP2D6 interindividual variability (all screening approaches). Only studies that reported comparisons of phenotypic variability / metabolism types, i.e., ultra-rapid, extensive, intermediate, and poor were included. All outcome measures reflecting drug efficacy (pharmacokinetics) and/or reduction in ADRs were eligible for inclusion. All study designs, including quantitative and qualitative but excluding reviews and meta-analyses, were eligible for inclusion.

### 2.2 Information Sources and Searches

The review comprised an electronic database search conducted using Web of Science (WoS) core collection (1970-2021), one of the largest and most authoritative multidisciplinary citation indexes of the most influential peer-reviewed scholarly journals, including those serving the field of biochemistry. Due to limited empirical studies in the area, no publication date restrictions were imposed to ensure that all relevant studies were identified. All searches were conducted between 17 January 2021 and 29 March 2021.

Search terms combined relevant keywords “CYP2D6” or “Cytochrome P450 2D6”, “opioid” or “opiate” and “dosage”, with standard Boolean operators (e.g., “AND”, “OR”) and truncations and wild card characters (e.g., “*”, “$”). Searches were conducted by entering the following terms:

| Search Terms |
| --- |
| • CYP2D6 [MeSH] in opioid* dosage*<br>• CYP2D6 [MeSH] in opiate* dosage*<br>• Cytochrome P450 2D6 [MeSH] in opioid* dosage*<br>• Cytochrome P450 2D6 [MeSH] in opiate* dosage* |

### 2.3 Study Selection

The study selection process is illustrated in Figure 1. All identified studies were screened against the eligibility criteria (see Table 4) and excluded at title, abstract, and full paper screening stages.

### 2.4 Data Abstraction

A single reviewer (RC) independently abstracted all data items using a custom data collection form developed to standardise the process (see Table 6). Data items abstracted were [author(s), date of publication], [population, sample size], study design, intervention (method of screening and opioid dosage), comparison (phenotypes), outcome measures (drug efficacy and ADRs), and study results.

**Table 6.** Main characteristics and data items of included studies

| Author(s)/Date | Population/Sample Size | Study Design | Intervention | Comparisons | Outcome Measures | Results |
| --- | --- | --- | --- | --- | --- | --- |
| Takahashi et al., (2017) (USA) | Primary care adult outpatients; 54% female, median age 49 years / N = 929 | Naturalistic observational cohort study (9 years) | Genotypic variability screening using xTAG CYP2D6 kit (Luminex Corp); medically prescribed opioid treatments (oxycodone, codeine, hydrocodone, tramadol, morphine, metoprolol, amitriptyline) | 7 phenotype metaboliser groups: ultrarapid, extensive to ultrarapid, extensive, intermediate to extensive, intermediate, poor to intermediate, poor | Hospital and emergency department (ED) admission for ADRs | Increased risk of ADR hospitalization for among ultrarapid metabolisers compared to all other phenotypes; increased risk of ADR ED visits among ultrarapid and poor phenotypes compared to all other phenotypes |
| Fonseca et al., (2011) (Spain) | Opioid dependent disorder patients (Caucasian) receiving a stable methadone dose for the last two months; 71% male; mean age 38 years / N = 105 | Clinical trial | CYP2D6 genotypic variability screening using deletion-specific long range PCR reaction; medically prescribed methadone (methadone maintenance treatment) | 3 phenotype metaboliser groups: ultrarapid, extensive, poor | (R)-, (S)- and (R,S)-methadone metabolic disposition (ng/ml) | Higher concentration of methadone in blood plasma of patients who encoded ultrarapid metaboliser compared to combined extensive/poor metabolisers; 347 ng/ml [SD = 279] vs. 265 ng/ml [SD = 269]) |
| Bastami et al., (2014) (Sweden) | Healthy volunteers; 9 males, 10 females; mean age 18 years / N = 19 | RCT | CYP2D6 genotypic variability screening conducted using NorDiag Arrow and gene-amplification PCR system 9700; medically prescribed oral tramadol | 3 phenotype metaboliser groups: extensive, intermediate, poor | Tramadol and O-desmethyiltramadol metabolic disposition (ng/g) $C_{max}$ & $t_{max}$ ADRs - drug related symptoms (DRS) | High positive correlation between functional alleles and the metabolic ratio of O-desmethyiltramadol, with the greatest rate of tramadol metabolism in extensive phenotype metabolisers |
| Wang et al., (2019) (China) | Laparoscopic patients; 18 females, 41 males; mean age 57 years / N = 59 | RCT | Genotypic variability screening (including CYP2D6) using TaKaRa PCR kit and QIAGEN Sequencing Reaction Kit; postoperative intravenous opioid treatment (Sufentanil, Oxycodone) | 4 phenotype metaboliser groups: ultrarapid, extensive, intermediate, poor; oxycodone vs. sufentanil groups | Postoperative ADRs, opioid dosage and VAS scores | No association between CYP2D6 polymorphism and postoperative ADRs, opioid dosage or opioid efficacy based on VAS scores |
| Jeong, et al., (2019) (Korea) | Healthy volunteers, Korean nationals; mean age 25 years / *N* = 23 | Modelling study (primary data) | CYP2D6 genotypic variability screening using deletion-specific long range PCR reaction; orally administered tramadol based on CYP2D6 phenotype | 3 phenotype metaboliser groups: extensive, intermediate, poor | Tramadol and O-desmethyiltramadol metabolic disposition | Developed models could be applied to predict concentration-dependent toxicities according to CYP2D6 genotype; only poor metabolisers reached the toxic range following 100mg administration of tramadol |

### 2.5 Risk of Bias and Quality Assessment

The CASP (Critical Appraisal Skills Programme, 2018) critical appraisal tool guided assessment of risk of bias and methodological quality of studies included in the review. Internal validity, reporting of results and conclusions and impact were the main areas appraised and reported as part of narrative synthesis.

### 2.6 Synthesis of Results

Data items from all eligible papers included in the final review were tabulated (see Table 6) and formed the basis of a qualitative synthesis and narrative analysis.

## 3. Results

### 3.1 Study Characteristics

One study was conducted in North America, two in Europe (Spain, Sweden) and two in East Asia (China, Korea). All studies used adult samples. Two of the studies used healthy volunteers and the remaining three studies used patient samples (post-surgical laparoscopy, primary care, opioid dependent). Nationality and ethnicity of the participants were explicitly reported in two of the studies: Korean nationals (Jeong et al., 2019), Caucasian (Fonseca et al., 2011). Sample size can be described as small (*N* < 30) in Bastami et al., 2014 and Jeong et al, 2019 medium (*N* = 59) in Wang et al., 2019 and large (*N* > 100) in Takahasi et al., 2017 and Fonseca et al., 2011.

Bastami et al., 2014 and Wang et al., 2019 used an RCT design, considered the ‘gold standard’ for study design (Hariton & Locascio, 2018). Other designs included clinical trial (Fonseca et al., 2011) and (Jeong at al., 2019), naturalistic cohort observation (Takahasi et al., 2017), and primary data modelling (Jeong et al., 2019). Whilst the inclusion of modelling studies in systematic reviews has been questioned, there is stronger support for those based on primary data, as is the case with the one modelling study included here.

All studies screened participants for CYP2D6 genotypic variation using polymerase chain reaction (PCR) based screening methods. Considered a reliable method for gene amplification and genotype analysis (Strom et al., 1994), PCR involves the synthesis of complementary DNA (amplicons) through the addition of nucleotides in the 3’ to 5’ direction assisted by a primer and DNA polymerase (see figure 6). Amplification of these genes can then be identified for their specific allelic makeup. Only one study stated the qualitative genotyping assay instrument used for CYP2D6 allele identification, xTAG kit. The remaining studies did not state the genotyping assay used, potentially making cross-study comparisons of phenotype groups problematic.

**Figure 6.**
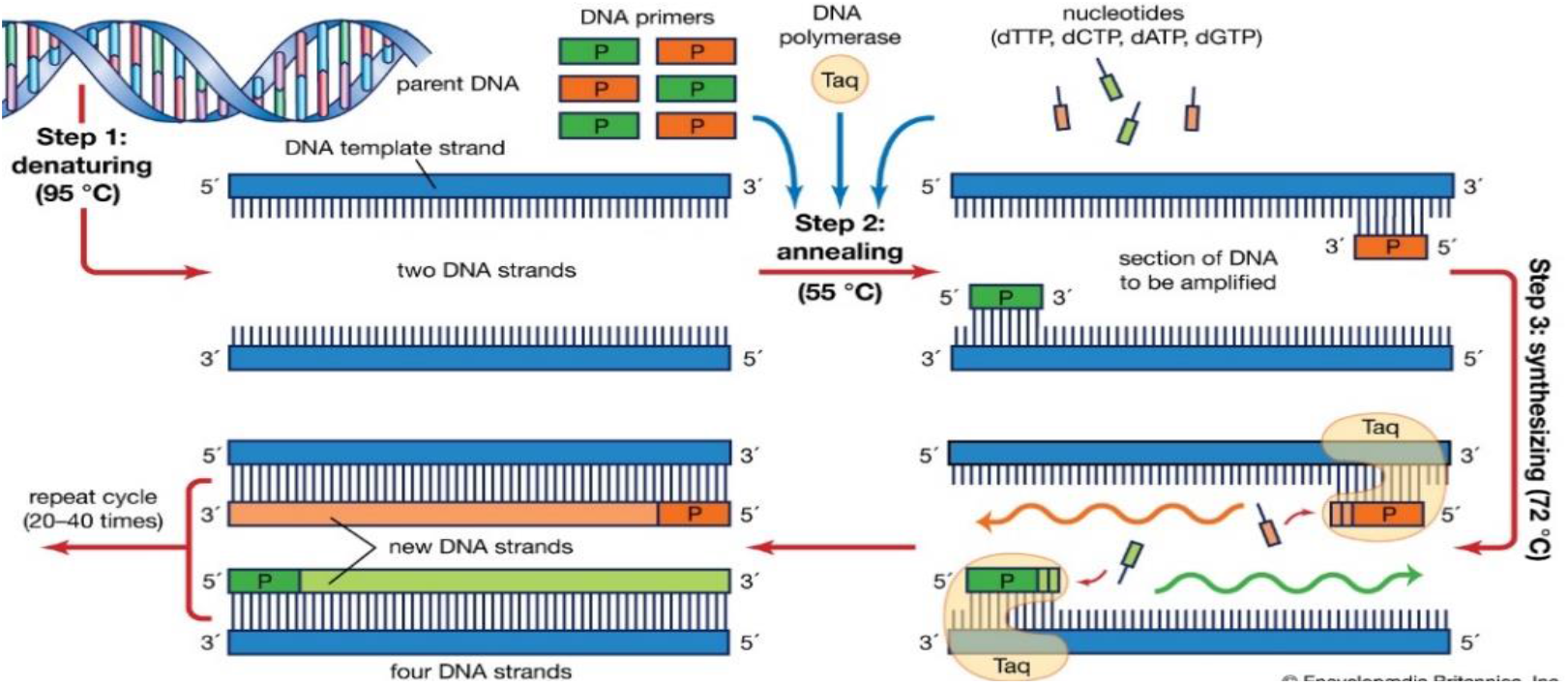
*Polymerase Chain Reaction* (Britannica, 2019)

All studies involved an analgesic-based pharmaceutical intervention. Three studies used a single opioid intervention (tramadol, n= two and methadone n= one). The two remaining studies used multiple opioids; oxycodone and sufentanil in one study, oxycodone, codeine, hydrocodone, tramadol, morphine, metoprolol, amitriptyline in the final study.

All studies performed comparisons between phenotypic groups. CYP2D6 phenotypes were determined using genotypic screening outcomes. The majority of studies (4/5) categorised phenotypes into three or four groups, UM, EM, IM and PM. The remaining study introduced three further groups spanning the existing phenotype categories, EM to UM, IM to EM, and PM to IM. Only one study also performed a comparison between opioid types, oxycodone vs. sufentanil.

Outcome measures adopted in all studies defined as phenotype-dependent changes in the pharmacokinetics of opioid analgesic activity (PD/PK modelling) and occurrence of ADRs. Three studies reported pharmacokinetic outcome measures, plasma concentration of methadone (Fonseca et al., 2011), plasma concentration of O-desmethyltramadol/tramadol (ODT/TRA) (Bastami et al., 2014) and (Jeong et al., 2019). Three studies reported ADRs outcome measures, hospital admission and ED visits for ADRs (Takahasi et al., 2017) and (Bastami et al., 2014) and postoperative ADRs/drug related symptoms (Wang et al., 2019). One study reported VAS scores as a measure of drug efficacy, but provided only a limited account of the measure, making interpretation of the results related to the VAS problematic.

### 3.2 Pharmacokinetics and CYP2D6

Three of the studies (Fonseca et al., 2011), (Bastami et al., 2014) and (Jeong et al., 2019) considered the implications of CYP2D6 interindividual variability on the pharmacokinetic activity of opioids through pharmacological trials. Two of the studies (Bastami et al., 2014) and (Jeong et al., 2019) adopted single or multiple-dose orally administered tramadol (TRA) as their principal pharmaceutical intervention considering the relationship between CYP2D6 polymorphs and the associated primary metabolite of tramadol, O-desmethyltramadol (ODT). Both studies adopted ODT/TRA metabolic disposition as the primary outcome measure to plot the relationship between CYP2D6 polymorphs and the metabolic activity of tramadol. Each study adopted similar statistical approaches and pharmacokinetic parameters, which included C_max_, t_max_ and AUC_0–10_, AUC_0–1_. Using these parameters to plot concentration-time graphs of ODT/TRA and conduct correlation and linear regression analysis for different phenotype metaboliser groups, Bastami et al., (2014) reported a significant association between CYP2D6 phenotype and both AUC MR (*p* < .01) and C_max_ ODT/TRA [ratio] in the blood plasma (MR) (*p* = .005). Mean values of AUC MR and C_max_ were highest in EM (0.41 ± 0.1 & 0.33 ± 0.1) followed by IM (0.24 ± 0.1 & 0.2 ± 0.1) and PM (0.09 & 0.07). The coefficient of determination (R^2^) represents the proportion of the variance in ODT/TRA explained by time after tramadol intake, indicating the capacity of time after intake to predict ODT/TRA [ratio]. Positive correlations between mean MR and time after tramadol intake were reported for EM (R^2^ = .97) and IM (R^2^ =. 96) but not PM, where MR remained constant across time after drug intake intervals. Expressed as rate of change (MR/time) derived from the linear regression equations reported by Bastami et al., phenotype-dependent differences in the linear increase in MR with time after tramadol intake are evident, EM = 0.312/1, IM = 0.0215/1, and PM = 0.0031/1. Based on their findings, Bastami et al. concluded that an ‘estimation of the time of drug intake using the MR of ODT/TRA is not valid without CYP2D6 genotyping’ (p. 132). Similar to Bastami et al., Jeong et al. used clinical trial primary data to develop a pharmacokinetic model predicting changes in time-concentration profiles for tramadol and ODT/TRA (plasma) in relation to CYP2D6 genotype.

Findings showed genotypes encoding PM resulted in ‘very low’ plasma concentration of ODT (mean C_max_ 0.643 ng/mL) compared to UM (mean C_max_ 126.8 ng/mL), EM (mean C_max_ 83.8 ng/mL) and IM (mean C_max_ 40.83 ng/mL) groups. Even at recommended tramadol dose (100mg), C_max_ of tramadol reached toxicity in PM, and in UM where the C_max_ of ODT exceeded the therapeutic window, leading Jeong et al. to conclude ‘that the frequency of concentration-related ADRs may be reduced by optimizing the dosing regimen according to CYP2D6 genotype’ (p. 617). The final study concerned with pharmacokinetics of opioids and CYP2D6 polymorphs, involved methadone as the pharmaceutical intervention. Fonseca et al., used plasmatic concentrations of (R, S)-, (R) and (S) - methadone to study the influence of allelic variants of genes encoding CYP450 enzymes, including CYP2D6, in opioid dependent patients undergoing methadone maintenance treatment (MMT). Only CYP2D6 phenotypes showed significant differences in methadone blood plasma concentration. Higher concentration of (R)-, (S)- and (R,S)-methadone in blood plasma were reported for patients encoding UM compared to all other phenotypes, with significant differences reported for UM vs. EM: (R)-methadone UM 1275 ng/ml [SD = 484] vs. EM 503 ng/ml [SD = 416], *p* = 0.002; (S)-methadone UM 707 ng/ml [SD = 267] vs. EM 263 ng/ml [SD = 207], *p* < 0.001; and (R,S)-methadone UM 568 ng/ml [SD = 262] vs. EM 239 ng/ml [SD = 256], *p* = 0.048. Furthermore, an ‘overrepresentation’ of UM phenotype in patients who responded to the MMT programme compared to patients who continued illicit drug use (7% vs. 0%, *p* = 0.032) was also reported. No dose restrictions were in place on the MMT programme. UM received significantly higher dose than EM (177 mg/day [SD 96] vs. 95 mg/day [SD 60], *p* = 0.043), and PM received marginally lower dose that other phenotypes (87 mg/day, SD 67). Fonseca et al. concluded that ‘pharmacokinetic factors could explain some but not all differences in MMT outcome and methadone dose requirements.’

Consistent findings across the three studies that focus on pharmacokinetics highlight the significance of CYP2D6 phenotypic variability against the concentration of active opioid metabolites (ODT and methadone) in the blood plasma. Taken together, these findings provide evidence supporting the significant role of CYP2D6 in the pharmacokinetics of both tramadol and methadone.

### 3.3 Adverse Drug Reactions and CYP2D6

The remaining two studies included in the review investigated the association between CYP2D6 phenotypic variability and opioid-based ADRs. In a large-scale longitudinal cohort study of primary healthcare patients, Takahashi et al., found an increased risk of both hospitalisation and emergency department (ED) visits for ADRs in individuals who encoded for UM phenotypes compared to all other phenotypes. Results showed 47% of UM were hospitalised compared to 30% of EM (*p* = 0.07). This increased risk was also present in the rate of ED visits, where 62% of individuals who encoded UM phenotype had an ED visit, compared to 49% of individuals who encoded EM phenotype. Based on their findings, Takahashi et al. conclude that ‘there may be clinical utility in preemptively genotyping patients to decrease health care use’ (pp. 45-46). Wang et al. (2019) recorded ADRs in postoperative patients according to the presence of the CYP2D6*10 allele, which encoded a decreased CYP2D6 activity. Genotypic frequency of occurrence of CYP2D6*10 was classified into CC, CT and TT in a randomised trial comparing oxycodone and sufentanil. ADRs were defined as nausea, vomiting, respiratory depression and dysphoria. Although some differences were apparent from the descriptive analysis presented, these did not reach statistical significance and an absence of observed differences in ADRs between CYP2D6*10 genotype groups was reported (*p* > 0.05). Furthermore, no significant differences in visual analogue pain scale (VAS) scores or total patient controlled analgesia (PCA) dosage (i.e., drug efficacy) were found between CYP2D6*10 genotypes (*p* > 0.05). Based on significant differences reported for other gene polymorphisms included in their study, Wang et al. conclude ‘gene polymorphism in patients undergoing laparoscopic gastric or intestinal cancer surgery is associated with opioid efficacy and adverse reactions’ (p. 9409). A third study included in the review, Bastami et al. (2014), focusses on pharmacokinetics but does also report drug related symptoms (DRS) against CYP2D6 phenotypes. DRS represented ADRs in the study and included nausea, vomiting, dry mouth, fatigue, headache and sweating. Similar to Wang et al., no association between tramadol DRS and CYP2D6 polymorphisms were found.

Differences in ADR outcome measures and genotyping approach adopted in the three studies should not be ignored and may account for the lack of consensus in the findings regarding the association between CYP2D6 genotypes and ADRs. Nevertheless, the evidence from these studies does not provide unequivocal support the role of CYP2D6 phenotypic variability in opioid induced ADRs.

### 3.4 Risk of Bias and Quality Assessment

Sample size (*N*) was small in two of the studies (Bastami et al., 2014 and Jeong et al., 2019) which was exacerbated by phenotypic grouping, where participants numbers (*n*) were as low as one in the PM group in both studies. This limited statistical analysis and confidence in some aspects of analysis. Some parts of statistical analysis and significance levels were not fully reported in some studies. Whilst all studies investigated phenotyping, establishing safe dosing was not the primary aim of all studies. With the exception of one modelling study (Jeong et al.), study design could be considered broadly homogeneous. Similarly, the degree of homogeneity in pharmacokinetic outcome measures was good, allowing comparison across studies. This was not the case for ADR outcome measures, which were varied and lacked full reliability, validity and self-reporting. The largest of the ADR studies, Takahashi et al., reported background analysis of relevant health behaviours such as smoking and alcohol consumption, with no significant differences between phenotypes, and adjusted statistical analysis for age and sex. This increased confidence in the study’s findings. Finally, where the study opted to screen for a single CYP2D6 allele, as in the case of Wang et al., comparisons were limited to phenotypes, which the allele encoded for, restricting interindividual phenotypic variability and cross-study comparisons.

## 4. Discussion

Opioid prescribing has increased sharply over the last decade (Schnell, 2019) and an associated increase in the incidence of opioid related ADRs in patients presents a global public health risk, with significant human and financial cost (Khalil & Huang, 2020). The incidence of ADRs highlights limitations in existing dosing strategies that rely largely on height, weight, age and subjective factors such as pain scores. The aim of the review was to systematically interrogate the available published literature to establish whether there was evidence to support the potential of clinical polymorphic screening of CYP2D6 as a significant factor in determining safe and effective opioid dosing strategies.

Three studies considered pharmacokinetics of opioids measured against CY2PD6 phenotypic variability. Using concertation-time graphs, the studies provided convincing evidence for the association between plasma concentration of active metabolites (methadone and O-desmethyltramadol) and CYP2D6 polymorphs. Congruent findings across the studies revealed that individuals encoding for UM/EM CYP2D6 phenotypes were exposed to the highest concentration of active opioid metabolites in their bloodstream compared to IM/PM. Varying rates of active metabolites is explained by the activation of opioid compounds via conjugative O-demethylation and N-demethylation reactions by CYP2D6. Dealkylation of opioids via CYP2D6 pathways results in an increased analgesic activity as a result of heightened interactions at the MOR and increased affinity for μ opioid receptors. The effects of CYP2D6 polymorphs on the pharmacokinetics of opioids, causing differences in the phase I activity of CYP2D6, are well established. The findings of the current review relating to pharmacokinetics are consistent with the known pharmacogenomics of CYP450s and the activity of CYP2D6 on analgesic compounds and support the potential significance of CYP2D6 screening in developing improved opioid dosing strategies.

Three studies compared CYP2D6 phenotypes against observed and self-reported ADRs. Findings across the studies were inconsistent. The largest study (Takahashi et al., 2017), with 929 participants, reported an increased risk of ADR related hospitalisation and ED visits in UM CYP2D6 phenotypes compared to all other phenotypes. The other two studies (Bastami et al., 2014; Wang et al., 2019), both using self-report and observed drug related symptoms such as vomiting and headaches as a measure of ADRs, reported no significant association between CYP2D6 polymorphisms and ADRs. Whilst the lack of consensus in findings relating to ADRs limits inferences regarding the significance of CYP2D6 screening in developing opioid dosing strategies, heterogeneity in ADR outcome measures and genotyping approach used in the three studies may explain the discrepancy.

Of the five studies included in the review, four reported an association between CYP2D6 phenotypic variability and opioid metabolising. On balance, the evidence reviewed and presented here supports the potential significance of CYP2D6 phenotyping in developing accurate opioid dosing strategies. However, despite outlining its pivotal role in the metabolic pathway of opioids, CYP2D6 may be limited by a lack of, yet proven, clinical significance. Executing a programme of CYP2D6 clinical screening will be costly but should be considered in light of the alternative human and financial cost of escalating opioid related ADRs.

### 4.1 Limitations

The review did not include studies published in languages other than English and limited resources meant that screening and data abstraction was conducted by a single reviewer. Although the review searched arguably the leading online database for relevant literature, Web of Science, multiple database searches may have yielded additional eligible studies. In an assessment of risk of bias and methodological quality, concerns were raised regarding sample size, outcome measures and statistical reporting. Small sample size in two studies restricted statistical analysis and confidence in the findings. Heterogeneity in ADR, but not pharmacokinetic outcomes measures, presented difficulties for cross-study comparisons and interpretation of findings. Concerns regarding the reliability and validity of observational and self-report ADR outcome measures were also noted. Furthermore, one study adopted a variation on genotyping approach that resulted in a restricted range of phenotypes not consistent with the other studies. This constituted a further obstacle for cross-study comparisons. However, level homogeneity in study design and pharmacokinetic outcomes measure were judged as good, reducing risk of bias.

## 5. Conclusion

In summary, the majority of studies included in the review (4/5) reported significant effects of CYP2D6 on opioid metabolising. The evidence was strongest in relation to CYP2D6/opioid pharmacokinetics. Lack of consensus in findings somewhat undermined the evidence regarding the association between CYP2D6 phenotypes and ADRs, although significant effects were reported. Based on the available evidence, it seems reasonable to conclude that CYP2D6 is a significant factor in opioid metabolising (i.e., *proof of concept*), that may be critical in determining safe and effective dosage. Faced with continued growth in opioid prescribing and inevitable rise in ADRs, research science must redouble efforts to ensure safe and effective opioid dosing strategies. That CYP2D6 metabolic activity along a large number of opioid pathways determines the analgesic activity by phase I dealkylation reactions, which result in varying affinity for the μ opioid receptors, suggests it as a principal candidate to help circumvent opioid-based ADRs.

The limited number of eligible studies identified in the review suggests the need for an increase in primary studies investigating CYP2D6. In general, these studies should be conducted with larger samples and adopt consistent screening/phenotyping approaches and outcome measures, with understanding the role of CYP2D6 in opioid activity for accurate dosage as the principal objective. Future studies should also strive to establish the clinical significance of CYP2D6 screening. Despite evidence outlining CYP2D6 as an important determinant in opioid metabolism by demethylation reactions and downstream ADRs as a result of these metabolic reactions, there is little research and development in the production of fast acting identification tools for high frequency CYP2D6 SNPs that could provide a cheap, scalable solution to the fast-evolving issue that is opioid-based ADRs. These tools could be further specified by high frequency geographically distributed alleles, as a solution for the scalability of these clinical screening tools. Doubtless, the relevance CYP2D6 in improving dosage strategies will involve an analysis of cost, where the development of ‘cheap’ methods of CYP2D6 phenotyping will prove pivotal. Large-scale longitudinal cohort studies will also be necessary to provide data on the impact of CYP2D6 phonotypes screening and its capacity to improve analgesia and mitigate opioid-based ADRs.

## Data Availability

All data can be made available for use upon request

## Notes

### Competing Interest Statement

The authors have declared no competing interest.

### Funding Statement

No funding provided

